# A Cluster-based Model of COVID-19 Transmission Dynamics

**DOI:** 10.1101/2021.06.02.21258243

**Authors:** B Shayak, Mohit M Sharma

## Abstract

Many countries have manifested COVID-19 trajectories where extended periods of constant and low daily case rate suddenly transition to epidemic waves of considerable severity with no correspondingly drastic relaxation in preventive measures. Such solutions are outside the scope of classical epidemiological models. Here we construct a deterministic, discrete-time, discrete-population mathematical model which can explain these non-classical phenomena. Our key hypothesis is that with partial preventive measures in place, viral transmission occurs primarily within small, closed groups of family members and friends, which we call *clusters*. Inter-cluster transmission is infrequent compared to intra-cluster transmission but it is the key to determining the course of the epidemic. If inter-cluster transmission is low enough, we see stable plateau solutions. Above a cutoff level however, such transmission can destabilize a plateau into a huge wave even though its contribution to the population-averaged spreading rate still remains small. We call this the *cryptogenic instability*. We also find that stochastic effects when case counts are very low may result in a temporary and artificial suppression of an instability; we call this the *critical mass effect*. Both these phenomena are absent from conventional infectious disease models and militate against the successful management of the epidemic.

## INTRODUCTION

### §1. Plateaus and waves

In many countries, the COVID-19 case trajectories have shown a “plateau” or constant and low daily case rate for an extended period. Oftentimes, these plateaus have given way to mountains of cases with little or no warning. India is an extreme example, where a second wave of abnormally high reproduction number (upto 2·1 in the worst affected cities) arose in April 2021 after months of declining daily case counts followed by weeks of steady and very low counts. (The reproduction number *R* is defined as the average number of people to whom one case transmits the disease.) In European countries like Germany, Italy and Slovakia, similar even if less dramatic scenes played out earlier in the pandemic’s history, while USA went through as many as four waves of epidemics during the past fifteen months. Very recently, Japan has been hit by a fourth wave threatening the Olympics while Taiwan has started exhibiting its first wave of the disease after more than a year of plateauing at nearly zero level.

Plateaus are outside the ambit of lumped parameter or compartmental infectious disease models such as S-I-R and S-E-I-R. A compartmental model based on delay differential equations (DDE) has recently been proposed by our group [1]. This model is more realistic and more versatile than S-E-I-R but a constant case rate is still not a generic solution; it occurs only if *R* equals exactly unity, which happens for specially chosen parameter combinations and infection levels. All these models (which we collectively term as “classical”) do have second waves corresponding to reopening but a major change in epidemic trajectory must necessarily be caused (and hence immediately preceded) by a major relaxation in the level of non-pharmaceutical interventions (NPI) like mobility restrictions or masking. This was not the situation in some countries.

In May 2020, Thurner et. al. [2] first found a constant case rate as a stable, generic solution of a disease transmission model. These authors have considered a network of people in the shape of a circle with every lattice point being connected to its nearest neighbours. They have then connected some of the nodes to additional nodes, located far away on the circle. This accounts for the fact that people tend to interact frequently with their families and much less frequently with outsiders. In other words, Thurner et. al. differ from classical models by accounting for heterogeneity in people’s interaction patterns. They find that, if the average degree *D* of the network remains below a critical value then the infection spreads at a constant rate before dying out, while if the average degree exceeds this value then the epidemiological curve resembles the bell-shaped or wave solution of ODE models. An approximate expression has been calculated for the critical degree *D*_*c*_ in terms of the transmission probability and the transmissibility interval.

While Ref. [2] appears to be a significant advance relative to the classical models, there are some overlaps with these models as well. For example, the nearest-neighbour links are interpreted as family ties while the long-distance bridge links are interpreted as “social contacts outside the local community (family)”. Thus, the constant-rate solution appears to hold only when there is a hard lockdown. Indeed, in a note added to the Supplement during proof (July 2020, by which time second waves had broken out in a lot of places), the authors mention that “many countries have (at least partially) taken back many NPIs, resulting in a (seemingly exponential) resurgence of daily infections ….. [which] is maybe not yet the ‘second wave’ but just the logical [fallout of the] reduction of social distancing and [consequent] increase of *D, d* [transmissibility duration] and *ε* [probability of a bridging link existing between two distant nodes]”. Indeed, the statement that in the network model, the epidemic is contained below a certain average network degree *D*_*c*_ and evolves naturally above it is substantively the same as the statement that in the DDE model [1], the epidemic is contained below a critical interaction rate (for which *R*_0_ = 1) and evolves naturally above it. The nature of the containment solution is different in the two models (constant rate in Ref. [2] vis-a-vis decaying rate in Ref. [1]), but containment and wave solutions are still separated by a change in a population-averaged parameter.

In February 2021, Tkachenko et. al. [3] conjectured that the plateaus and waves are generated as a result of temporal heterogeneity in interaction rate, rather than inherent heterogeneity in our interaction patterns. Temporal variation refers to the fact that a person will have a high interaction rate with others on some days, for example when s/he attends a party, and a lower interaction rate on other days, for example when s/he stays at home. To some degree at least, we would expect such fluctuations to get smoothed out when the case counts are high, and indeed their final model equation is quite similar to the conventional S-I-R model. Nielsen et. al. [4] have focussed on heterogeneity in people’s infectivity i.e. superspreading incidents. Very recently, Manrubia and Zanette [5] have made the contention that plateau solutions (*R* = 1) are primarily the result of individuals’ risk-averse behaviour rather than a heterogeneity effect.

We believe that heterogeneity in interaction patterns remains the most plausible hypothesis behind the non-classical epidemic trajectories. After all, most or all people socialize within narrow groups of family and friends – interactions outside this set are considerably rare. Therefore, we build a transmission model which takes this as the starting premise. Our model is ultimately deterministic although it is based on probabilistic concepts. We find that it is capable of predicting plateaus and waves as well as unexpected transitions from the first state to the second.

## MATHEMATICAL MODEL

### §2. No vaccination

Before starting the model development, we mention that in this entire Article, we shall ignore vaccination. This is because COVID-19 plateaus and waves occurred in most countries when zero or small fractions of their populations were vaccinated (Chile, using vaccines made in a certain country, is a notable exception). American and European plateaus and waves occurred in mid-late 2020 when there were no vaccines against the disease. India’s second wave started when only about 10 percent of the population had got their first dose. Other countries currently suffering from waves, such as Taiwan and Japan, are also lagging behind on vaccination. Therefore we leave this intervention for a future study.

### §3. Qualitative understanding

It is our observation that in many regions of India and USA, high levels of socioeconomic activity could be sustained for weeks on end while cases remained on a plateau – restaurants, cinema halls, places of worship as well as public transportation remained open without triggering case explosions. Similar phenomena must have occurred in other countries as well. We first argue how this is even possible. Masking is literally impossible in eateries, and we are aware that some violations have also been occurring in cinema halls, public transportation and other places. We hypothesize as follows. Firstly, the vast majority of symptomatic people go into quarantine or at least refrain from interacting in society, if for no other reason than that public sneezing or coughing makes one an immediate target of suspicion. Hence, almost all transmissions occur from asymptomatic or latent (pre-symptomatic) cases via speech and breathing. Masks render these mechanisms almost incapable of transmission [6], so the majority of spreading occurs in unmasked settings. Even there however, breathing and speech carry the infected droplets over a relatively short distance (speech farther than breathing). Thus, if the virus is present in a restaurant, it is very much likelier to spread among people seated at the same table than to jump from one table to the next (or to saturate the restaurant and infect everyone inside). Similarly, mask fault in a cinema hall might also infect only the nearest neighbour and not someone seated far away. In summary, our first hypothesis is that the overwhelming majority of transmissions occurs during close, unmasked interactions.

It is logical that a person will indulge in such interactions only with his/her family members and friends and not with strangers. Thus, even if A goes to 20 restaurants in two months, it is likely that her dining companions on all these occasions will be a subset of B, C, D and E. Similarly, B’s companions might be A and D as well as F while C’s might be all the previous plus G. Our second hypothesis comes now. We posit that, starting from a random person, if we keep extending these links of close contacts, then it is extremely unlikely that the chain will continue all the way upto the country’s last citizen. On the contrary, sooner rather than later, the links will cover no new person and the cycle will close. This will give us a group of social contacts with dense links amongst each other and no (or very few) links outside. We call this group a **cluster**. At a small office for example, all employees might belong to a single cluster; at a large company, there will be multiple clusters with the people in each cluster possibly belonging to the same rank or payscale, or sharing the same office space. By the nature of clusters, when the virus enters a cluster it will spread rapidly among its members, but will find it much more difficult to infect someone outside the cluster.

The two hypotheses – primary spreading from close interactions and these being confined within clusters – can explain why it is possible to have low disease prevalence even with public transport, entertainment venues and places of worship open at full tilt. At recreation venues we interact primarily within our cluster and facilitate intra-cluster transmission. As regards places of worship, we go there either alone or with family, pray and come back. Inside a holy place we do not spend time socializing with all and sundry. Finally, in public transit as well, we by and large keep to ourselves, or interact with members of our cluster if we are going together to an entertainment venue. Thus, case counts remain low all through these activities as the virus remains primarily confined within a few clusters.

Of course, all transmission cannot be occurring inside the clusters; if that were the case then the epidemic would not perpetuate. So we must now examine the mechanisms by which the disease can jump from cluster to cluster. There are two ways this can happen. The first is *unintentional* – during necessary activities like shopping or working, an unknowing case’s mask might happen to slip off just when a potential target is close by, or we may touch a contaminated surface/object and then our face, or the virus might just jump across a pair of masks etc. The second mechanism of inter-cluster jumping is social occasions where we *deliberately* interact outside of our cluster. For example, the invitees at a marriage gathering might include multiple families who hate each other and normally do not meet (and even less so in pandemic times). Similarly, a birthday bash hosted by a well-to-do IT sector company employee might feature half the staff in attendance, an occurrence which would not take place at a casual entertainment venue. At events like this, adherence to COVID-appropriate behaviour tends to be low; worse still, social norms require us to actively interact with many people present at the gathering and not just with our family members or close friends. Thus, a case present at such a gathering will likely transmit the disease to several others not belonging to his/her cluster.

For the purposes of model-building we need to distinguish between household transmission and transmission among friends. In the former situation, due to constant contact, a case will spread the disease to all household members as soon as s/he turns infectious. In the latter situation however, it will take a finite time to fully infect a group – for example, the at large case A might dine with B and C on one day and infect them both, C will fall sick next week and go to a movie with E so on; it is unlikely that everyone from A to Z will be simultaneously present at an eatery or movie theatre and get infected in one fell swoop. To achieve this distinction, we must make two assumptions. Firstly, we decouple households and clusters, taking the latter to include only those close contacts with whom a person does not live together. Secondly, we assume that, per household, there is exactly one member who is socially active i.e. part of a cluster, while the other members are completely cautious. For example, in a family where a young working woman lives with her retired parents, the latter two might not step out of the house during the pandemic while the woman goes to work and does the shopping etc. A cautious family member can also be a person who is not socially inactive but rigorously adheres to COVID-appropriate behaviour all the time. In either case, the only way that the cautious person catches corona is if the active person catches it first; the active one can catch it through intra-or inter-cluster transmission.

In view of the above, we build our mathematical model taking into account a four-tiered viral transmission process, as follows.

- **Household transmission :** As soon as one member of a household contracts the virus, all others immediately follow suit.
- **Cluster transmission :** When one member of a cluster gets infected, the others also gradually fall sick. The rate at which this happens and the fraction of people in the cluster who contract the infection are determined by the properties of the virus and the interaction rate of the cluster members among each other.
- **Unintentional cluster transition (UCT) :** Note that this is cluster *transition* and not *transmission* – a jump from one cluster to another. These are events like the accidental mask slippage in a shop mentioned above. In this category we also include the events where a person makes a new acquaintance outside his/her cluster and starts socializing with him/her – we expect that events of this kind will be rare overall.
- **Socializing external to cluster (SEC) :** These are events like the wedding or party mentioned above. Organized gatherings where people from different clusters interact with each other also belong to this category.

Intuition says that SEC events might be very dangerous from a public health perspective, since they are almost designed to facilitate large-scale inter-cluster transmission. We shall now verify this intuition through the mathematical model.

### §4. Quantitative model development

Our model treats time to be discrete and measured in days i.e. we construct a map rather than a flow. The population is also discrete as in an agent-based model, rather than continuous as in an ODE/DDE model. However, our model is ultimately deterministic rather than stochastic. Let us consider a city (see later for more clarification) of total population *N*, all of whom are initially susceptible. Let them belong to *N*1 households where *h* = *N*/*N*_1_ is the average household size. With the assumptions of § 3, there are *N*_1_ people who belong to social clusters and can contract the virus directly; once this happens, each of them transmits to the *h* − 1 other members of their household (or family). Thus we can focus on the disease dynamics only among the *N*_1_ socially active people, and add on the family cases at the end. The values we have chosen are *N* = 3,02,400 and *h* = 3 so that *N*_1_ = 1,00,800. Our *N* is chosen to enable a comparison with the results in Ref. [1] where we have used Notional Cities of a similar population, while the average household size of three is reasonable.

In the next step, we divide the *N*_1_ socially active people into *N*_*C*_ clusters. Here, we assume that all clusters have the same size *s*, so that *N*_1_ = *sN*_*C*_. The choice of 1,00,800 for *N*_1_ rather than exactly 10^5^ ensures that *N*_1_ is divisible by every number upto 30 except two-digit primes; this enables easy variation of *s*. The value we have gone with is *s* = 24, so that *N*_*C*_ = 4200. To fix the intra-cluster dynamics, we now need some elementary characteristics of viral transmission. We assume that the serial interval is 5 days [7,8] i.e. 5 days elapse between a primary case and a secondary case’s turning transmissible. We also assume that, once a person turns transmissible, s/he spends 3 days at large before recovering. The 3 day transmission interval is an average of the weeklong asymptomatic period and the approximately 1 day latent infectious period before a symptomatic case seeks quarantine [9]. The assumption of recovery after three days (rather than isolation, hospitalization or death) is simply for analytical tractability.

Our next necessary piece of information is that the basic reproduction number *R*_0_ of COVID-19 is somewhere between 2 and 5, depending on the viral strain etc [10,11]. This means that in the absence of interventions, one person spreads the disease to between 2 and 5 people. Assuming a value of approximately 2·5 for intra-cluster transmission, we find that 5 days (serial interval) after the first person is exposed, s/he infects 2·5 others, 5 more days later these infect 6·25 further people and so on. The reproduction number then starts decreasing rapidly as more and more people in the cluster turn immune. Considerations like this (together with a fair amount of heuristics) cause us to define the **cluster sequence** as [1; 3; 6; 7; 5; 1]. This means that 5 days after the virus is seeded into a particular cluster (i.e. the first person in the cluster becomes exposed), that cluster develops one case, 10 days after seeding it develops 3 more cases, 15 days after seeding it develops 6 additional cases and so on until 30 days after seeding it develops its final case. One person among the 24 is left immune. We discuss the effects of changing the cluster sequence in § 13.

The existence of a cluster sequence means that we can define each cluster to be in one of two states : susceptible if all members of the cluster are susceptible, and insusceptible as soon as the first member has been exposed, and for ever after (we assume permanent immunity, which seems to be valid so far [12,13]). During the 30 days it takes for the 23 cluster members to be infected, we treat the cluster as a whole to be insusceptible – we assume that any further exposure of cluster members during this period does not change the intra-cluster infection pattern.

At this point we can introduce the variables in the model. We count a person as a case on the day that s/he first turns transmissible. Let *y*_*i*_ be the cumulative number of cases occurred upto and excluding day #*i* and let Δ*y*_*i*_ be the additional cases occurring on day #*i* itself, so that *y*_*i*+1_ = *y*_*i*_ + Δ*y*_*i*_. Similarly, let *z*_*i*_ be the cumulative and Δ*z*_*i*_ the daily count of clusters which are seeded upto and on day #*i* respectively. *y* and *z* are obviously not independent; by definition of the cluster sequence if Δ*z*_100_ = 1 then Δ*y*_105_ gets a contribution of 1, Δ*y*_110_ gets a contribution of 3, Δ*y*_115_ gets a contribution of 6 and so on. We say “gets a contribution of” rather than “equals” because Δ*y*_105_ will also carry contributions from clusters which have been seeded on days #75, #80, #85, #90, #95 and #100. In addition we have the near-dummy variables for family cases *f*_*i*_ and Δ*f*_*i*_; since every active member will infect his/her two household members as soon as s/he turns infectious, and since the incubation period is 5 days, we have Δ*f*_*i*+5_ = 2Δ*y*_*i*_ and *f*_*i*+5_ = 2*y*_*i*_.

Having accounted for the first two transmission mechanisms of § 3, we now start work on the latter two i.e. the UCT and SEC modes. Our ultimate question is : given the case histories upto day #*i* and the interaction patterns for UCT and SEC, what is the number of susceptible clusters which get seeded on day #*i* ? This number will enable us to move one step forward in time i.e. from day #*i* to day #*i*+1. Since the phenomena involved are probabilistic but the model is deterministic, we must calculate the expectation value. We now demonstrate how to do this. Our baseline premises are : (*a*) on any given day all the *N*_1_ active people are equally likely to participate in UCT and SEC events, and (*b*) at both these events, clusters are mixed at random i.e. a case from cluster #101 is as likely to infect a member of cluster #102 as a member of cluster #2302. Assumption (*b*) is the equivalent of homogeneous mixing in classical epidemiological models but at the cluster rather than the individual level. As an aside, these assumptions ensure that the most realistic domain of validity of our model is a city but not a larger region, such as a state or country (or the world). A marriage function or birthday bash held in one city is likely to have maximum guests from the same city, and few if any guests from another city in the same state, another state in the same country, or another country. Aside over, since all cases remain at large for three days, the total number of at large cases present on day #*i* is *α* = Δ*y*_*i*−1_ + Δ*y*_*i*−2_ + Δ*y*_*i*−3_. SEC transmission is conceptually more concrete than UCT so we take that first. Let *n*_*S*_ (constant in time) be the total number of people who participate in SEC events every day – it does not matter whether it is a single gathering of size *n*_*S*_ people or fifty separate gatherings adding up to total *n*_*S*_ people which are taking place. We assume that if everyone is susceptible then one case attending an SEC event spreads the disease to *m*_*S*_ people at the event. We use the value *m*_*S*_ = 2.

The number of cases attending SEC events on day #*i* can be anything between 0 and *α*. The probability that the number is exactly *k* is

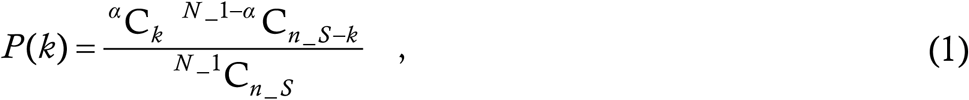

where the combination function ^*n*^C_*r*_ denotes the number of ways of selecting a team of *r* players from a pool of size *n*, and the underscores denote subscript when already in a super-or subscript position. The numerator in (1) consists of the number of ways of choosing *k* cases from the total *α* cases at large multiplied by the number of ways of choosing *n*_*S*_ − *k* healthy people from the *N*_1_ − *α* healthy people; the denominator is the total number of ways of choosing *n*_*S*_ people from *N*_1_ people.

Given that there are *k* cases participating in SEC events, by the model assumptions they transmit an infective dose of virus to *β* = 2*k* people (we say “transmit an infective dose” rather than “transmit the disease” since the recipient of the viral dose contracts the infection if and only if s/he is susceptible). The next question is, how many distinct clusters do these *β* people belong to ? This is relevant because if five recipients of infective viral dose belong to five different susceptible clusters then there will be five cluster sequences manifest over the next 30 days while if all the recipients happen to belong to the same susceptible cluster then (by the model assumptions) there will be only one cluster sequence manifest during this period. The possible number of clusters can range from between ⌈*β* / 24⌉ (smallest integer greater than or equal to *β*/24) and *β*, and we must ask what is the probability that the *β* people belong to exactly *b* clusters. With one caveat, this problem is identical to the following problem : there are *N*_*C*_ boxes and *β* balls; if any ball can go into any box, what is the probability that exactly *b* boxes contain at least one ball ? The caveat is that in the balls and boxes problem, a single box might contain all the balls while in the cluster problem, one cluster cannot contain more than 24 people.

We ignore the caveat for the following reason : in a typical situation, *N*_*C*_ >> *β,b* and there will be a very large probability that *β* people will belong to *β* or nearly *β* clusters. The spurious occurrences which we will pick up by ignoring the caveat (*β* people belonging to less than ⌈ *β* / 24 ⌉ clusters) will be extremely improbable anyway and the convenience gained will amply recompense the losses incurred. The caveat surmounted, the balls and boxes problem happens to be solvable in closed form. The numerator of the probability must be the number of ways of putting *β* balls into exactly *b* boxes (*b* ≤ *β*), while the denominator must include all possible ways of putting the balls into the boxes. For the numerator, we first select *b* boxes among the *N*_*C*_ ones, which can be done in ^*N*_*C*^C_*b*_ ways. Given the box selection, we must find the number of ways to express *β* as a sum of *b* natural numbers, including different orderings – this is the restricted composition function 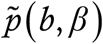 . The composition and not the partition function *p*(*b,β*) is relevant for the following reason : suppose we have 10 boxes in total and want to distribute 5 balls among 3 boxes, and suppose we have already selected the boxes number 2, 4 and 7. Then, it makes a big difference whether there are three balls in box 4 and one each in boxes 2 and 7 or three balls in box 2 and one each in boxes 4 and 7. The composition function 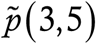 treats 3+1+1 as different from 1+3+1 and accounts for this difference while the partition *p*(3,5) treats the two as identical and fails to account for it.

The composition 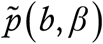 actually has a simple analytical formula unlike the partition. We can view a composition of *β* into *b* parts as introducing *b* −1 slats in between *β* beads arranged in a row on an abacus. So long as there is maximum one slat between two beads and there are no slats to the left or right of the row as a whole, the slats split the beads into exactly *b* parts – each different arrangement of slats corresponds to a different way of writing *β* as a sum of *b* numbers. In total, there are *β* −1 gaps between the beads which need to be filled by *b* −1 slats; the number of ways of doing this is ^*β*−1^C_*b*−1_. Thus, 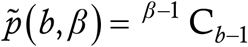.

To calculate the denominator of the balls and boxes probability, we imagine the *N*_*C*_ boxes lying in a row and focus on the walls separating one box from the next. The external walls of the leftmost and rightmost box must remain intact; the remaining *N*_*C*_ −1 walls plus the *β* balls can be arranged in a line in any order whatsoever. For this, among the total *N*_*C*_ −1+*β* positions which can be filled by either ball or wall, select *β* to fill with balls; the remainder automatically get filled with walls. The number of ways of doing this selection is ^*N*_*C*−1+*β*^C_*β*_. Putting all this together, and switching back from balls and boxes to viral recipients and clusters, the probability that *β* recipients belong to *b* clusters is

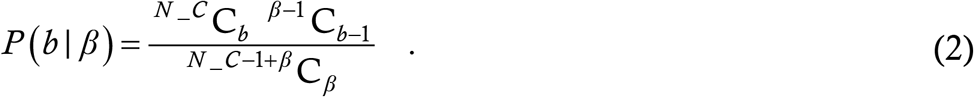

There is one more conditional probability to be taken care of. We know that *b* clusters are infected at SEC events, but how many of them are actually susceptible ? As the disease progresses, a higher and higher number of the *b* clusters will actually be insusceptible ones. On day #*i* there are *z*_*i*_ insusceptible clusters and *N*_*C*_ − *z*_*i*_ susceptible ones. The probability that among *b* randomly selected clusters, exactly *j* are susceptible is calculated just like *P* (*k*) in (1); we have

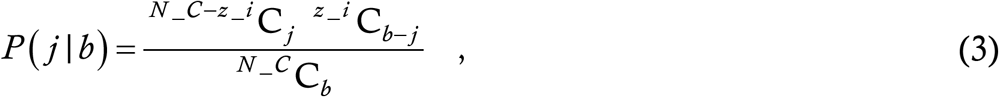

and all the conditional probabilities are on the table.

Now, the quantity of interest is the expectation value of the number of new susceptible clusters seeded on day #*i*. This can be calculated as

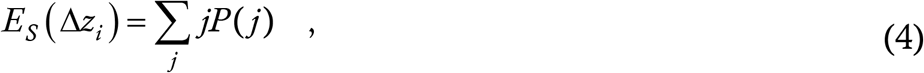

where the subscript *S* reminds us that this is the expected number of clusters seeded during SEC events and *P* (*j*) is the total probability that *j* susceptible clusters are seeded on day #*i*. So far we have the partial probabilities, • conditional *P* (*j*| *b*) : given that an infective dose of virus is introduced into *b* clusters, the probability that *j* of them are susceptible, • conditional *P* (*b*| *β*) [or equivalently *P* (*b*| *k*) since *β* = 2*k*] : given that 2*k* people receive infective viral doses, the probability that they belong to exactly *b* clusters, and • absolute *P* (*k*) : the probability that *k* cases are actually participating in SEC events on the day in question. Thus, given a pair *k,b* the probability of *j* susceptible clusters’ being seeded is *P* (*j*| *k,b*) = *P* (*k*) *P* (*b*| *k*) *P* (*j*| *b*). The total probability *P* (*j*) will be this summed over all possible *k* and *b. k* runs from 1 to a maximum of *α* while *b* runs from 1 to a maximum of *β*. Thus we have

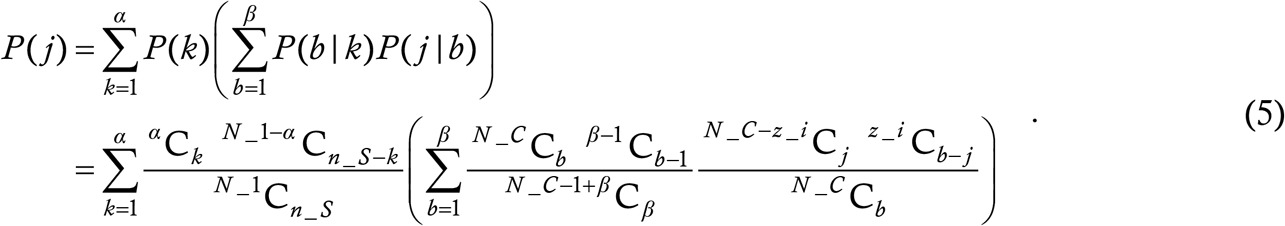

Now, we must implement the sum (4). For each *b, j* can run from 1 to *b* and we can pull the summation over *j* inside the second of the two sums in the above right hand side. Doing so gives us the expectation value

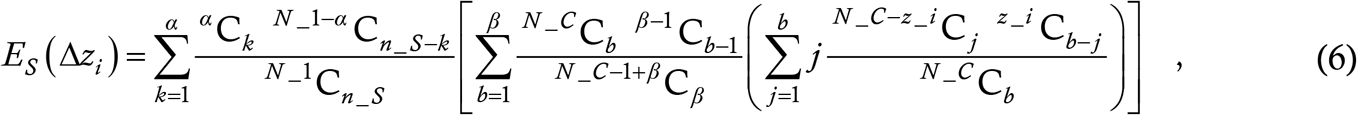

and the contribution of SEC events has been determined.

In a similar manner, we can calculate the contribution of UCT events to inter-cluster spread. Let *n*_*U*_ be the number of people participating in UCT events every day – these are the people who visit crowded markets, travel on crowded buses and trains etc. Let a case present at an UCT event transmit an infective dose of virus to *m*_*U*_ targets. Unlike for SEC, we expect that on the average *m*_*U*_ will be less than unity, in which case we can also interpret *m*_*U*_ as the probability *P*_*U*_ that a case successfully transmits an infective dose to a target at an UCT event. Since this probability is a parameter present in classical epidemic models as well, we use *P*_*U*_ rather than *m*_*U*_. However, we treat it just like *m*_*S*_, and keep open the possibility that the parameter value might exceed unity (in which case *m*_*U*_ is the only interpretation which makes sense). Then, we can repeat the argument for the SEC contribution. Given that there are *k* cases at large on day #*i*, the expected number of people who receive an infective dose is *kP*_*U*_. This is the equivalent of *β*, with one difference; while *β* = 2*k* was an integer by definition, *kP*_*U*_ will in general not be one, and the entire calculation is cast in terms of integers. To tackle this, we introduce the **roundoff function**. In this function, we define *γ* to be the integer nearest to *kP*_*U*_; since this can cause accumulating errors with increasing *i*, we also retain the difference between *γ* and *kP*_*U*_ and keep incrementing it with every successive *i*. When this difference exceeds +1 or −1, we include the correction to *γ*. This ensures that rounding errors do not accumulate in time. Roundoff apart, everything else is the same. Given that *γ* people have received an infective dose, we can calculate the probability that they belong to *b* different clusters and then that *j* of them are susceptible, and arrive at

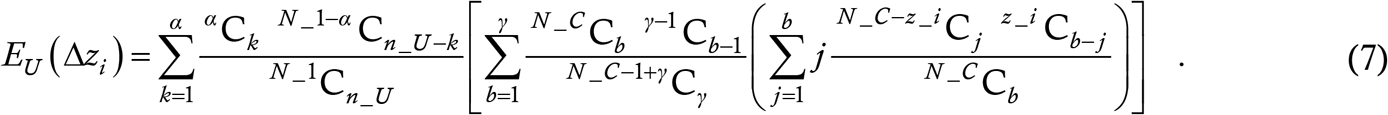

The expected total number of susceptible clusters seeded through SEC and UCT events is

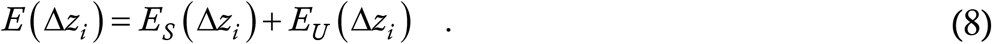

Since this will in general be a fraction, we again use the rounding off procedure described above, approximating it to the nearest integer and retaining, adding and correcting the error. Thus a roundoff on (8) gives Δ*z*_*i*_. At once we can implement Δ*z*_*i*_ times the cluster sequence over the next 30 days. Finally, we do *y*_*i*+1_ = *y*_*i*_ + Δ*y*_*i*_ and *z*_*i*+1_ = *z*_*i*_ + Δ*z*_*i*_ to complete one iteration of the map and move from day #*i* to day #*i*+1.

This is the detailed procedure which we follow; we now present it in the form of the algorithm which the computer is made to run.

### §5. The Algorithm

For those who skipped § 4, we quickly recall that our model is deterministic but discrete in time as well as in population. We recapitulate the variables and parameters and their definitions in the below Table; we also give the parameter values corresponding to a default or baseline solution.

The default initial condition is that eight clusters are seeded on the first day. We are now ready to present the algorithm itself.

#### Algorithm 1

The actual mathematical model as implemented on the computer.

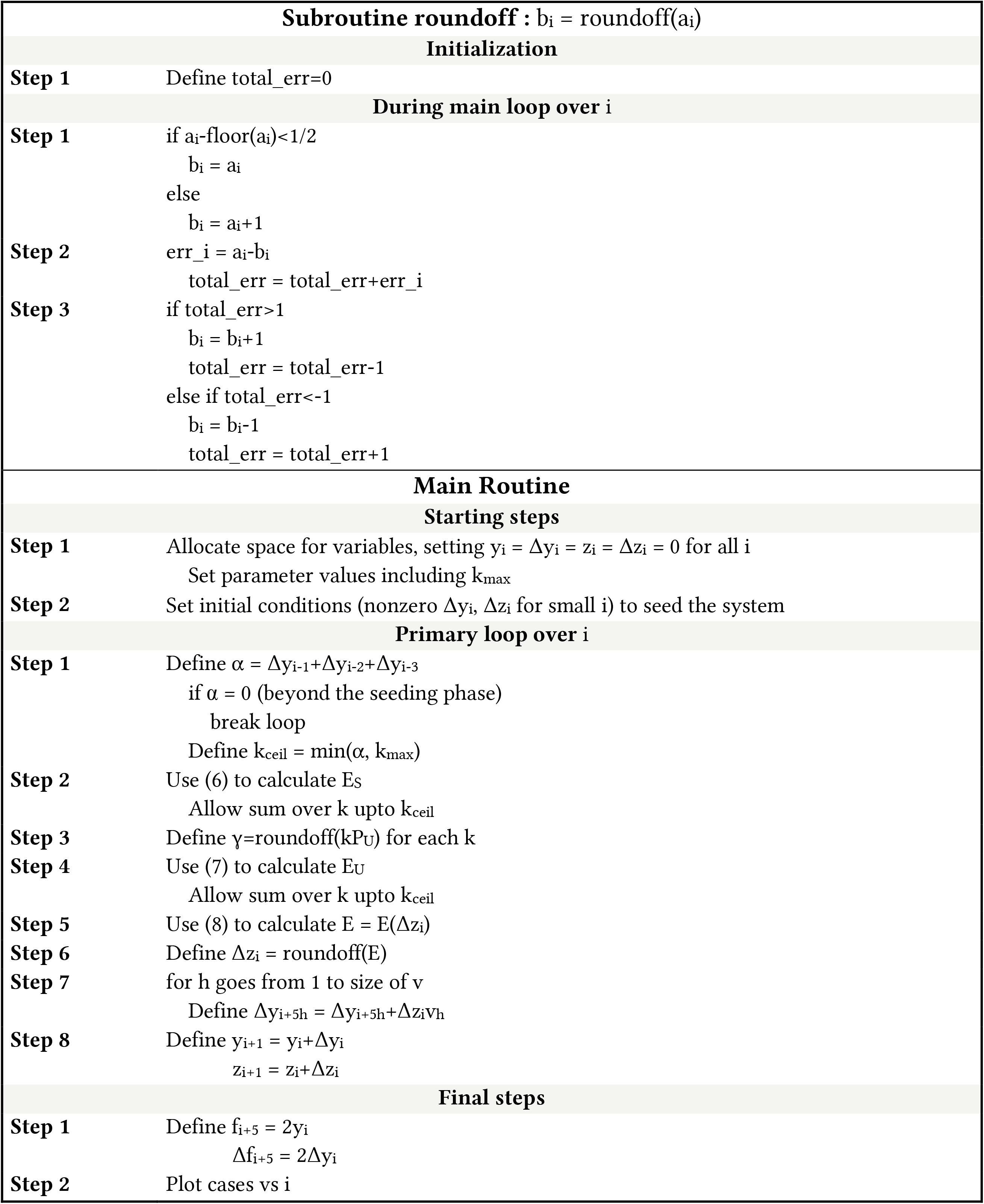

The parameter *k*_max_ in the algorithm does not appear in § 4. This is because it is introduced only for the sake of computational convenience. The sum over *k* in (6) and (7) should ideally run all the way upto *α*. However, with *n*_*U*_ and *n*_*S*_ both being significantly smaller than *N*_1_ (typically 1-10 percent), the probability that *α* or nearly *α* cases will all be participating in UCT or SEC events on the same day will be minuscule. Hence, we can define a cutoff *k*_max_ above which we shall just treat this probability to be zero, and ignore the error. The computational time and effort increase very rapidly with increasing *k*_max_ so it is worthwhile to choose its value judiciously. For all display results here, we choose *k*_max_ = 80.

Finally, the expressions (6) and (7) are next to impossible for the computer to handle unless the probabilities are inputted in a special manner; we shall describe this in the Appendix.

## SIMULATION RESULTS

### §6. The default solution

This corresponds to the solution of the model with the default parameter values from Table 1. We call these values defaults not because they have been obtained from any data fits etc but because variation of each parameter on either side of the default leads to different kinds of interesting behaviour. The time trace of the epidemic with these values is below. In all these plots, we shall show the cumulative case count as a blue line associated with the right hand *y*-axis and the daily case rate as grey bars associated with the left hand *y*-axis.

**Table 1:** Variables and parameters in the model. By “const.” we mean that the parameter value is kept unchanged in all simulation runs in this Article.

| Variable | Significance |
| --- | --- |
| $y_i$ | Cumulative number of socially active cases upto and excluding day $\#i$ |
| $\Delta y_i$ | Number of new socially active cases cropping up on day $\#i$ |
| $z_i$ | Cumulative number of insusceptible clusters upto and excluding day $\#i$ |
| $\Delta z_i$ | Number of new clusters turning insusceptible on day $\#i$ |
| $f_i$ | Cumulative number of cautious household cases upto and excluding day $\#i$ |
| $\Delta f_i$ | Number of new cautious household cases cropping up on day $\#i$ |

| Parameter | Significance | Default value |
| --- | --- | --- |
| $N$ | Total population | 3,02,400 (const.) |
| $h$ | Household size | 3 (const.) |
| $N_1$ | Socially active population | 1,00,800 (const.) |
| $N_C$ | Number of clusters | 4200 (const.) |
| $s$ | Size of the cluster | 24 (const.) |
| $\mathbf{v}$ | Cluster sequence | [1; 3; 6; 7; 5; 1] (const.) |
| $T$ | Serial interval | 5 days (const.) |
| $n_U$ | Number of people participating daily in UCT events | 10,000 |
| $P_U$ | Probability of transmission in a UCT event | 0.15 |
| $n_S$ | Number of people participating daily in SEC events | 0 |
| $m_S$ | Number of people infected by one case at a SEC event | 2 (const.) |

We can see that the epidemic continues for a long time at almost constant daily case rate before eventually dying out. This is the plateau solution.

### §7. Variation of *P*_*U*_

Here we present the case trajectories as *P*_*U*_ is varied. Keeping all other parameters fixed at their default values, we consider three representative values of *P*_*U*_ and display the time trace of the epidemic in the three panels of the below Figure. We have used the same *x*-and *y*-axis scalings in all panels so that the contrasts may be visually apparent.

For the lowest value we can see that the rate decreases monotonically starting from approximately the 100^th^ day, until the epidemic is eliminated by 250 days. This transitions to the plateau solution at approximately *P*_*U*_ = 0·135. This plateau is stable upto about *P*_*U*_ = 0·165 (this includes the middle panel of Fig. 2), after which it yields to a wave-like solution seen in the bottom panel. The behaviour obtained by increasing *n*_*U*_ while keeping *P*_*U*_ constant is remarkably similar, and we do not repeat the figures here.

### §8. Variation of *n*_*S*_

In this Section, the quantity which we vary is the number *n*_*S*_ of people participating in SEC events every day. Just as in Fig. 2, we present three plots of case trajectories for different values of *n*_*S*_ while all other parameters remain at their default values. Again, we use constant scalings on the axes across panels, to facilitate visual comparison. The scalings are different from the previous Section though.

In the last of the three scenarios, the epidemic progresses to what is conventionally known as herd immunity.

### §9. Effect of initial conditions (IC)

The discrete-population nature of the model means that the dependence of solutions on initial conditions is also non-trivial. In particular, a sufficiently large seeding caseload is required for the epidemic trajectory to be manifest. For example, if the default solution of Fig. 1 is seeded with four clusters instead of eight, then the epidemic terminates almost immediately instead of continuing at constant case rate. A more dramatic example is shown in the below Figure. Here, we choose *n*_*S*_ = 1000 and stick to the other default parameter values. Extrapolating from Fig. 3, this corresponds to a highly dangerous mode of operation and is expected to cause a huge wave. Instead of starting by seeding eight clusters however, this time we introduce some numbers of external cases Δ*y*_*i*_ on days 7, 10 and 13. These external cases are not part of any cluster but they participate in UCT and SEC events and spread the infection to the local cluster members. The panels of the Figure are titled by the vector [Δ*y*_7_; Δ*y*_10_; Δ*y*_13_]. Note that the axis scalings in the three panels this time are NOT the same !

**Figure 1:**
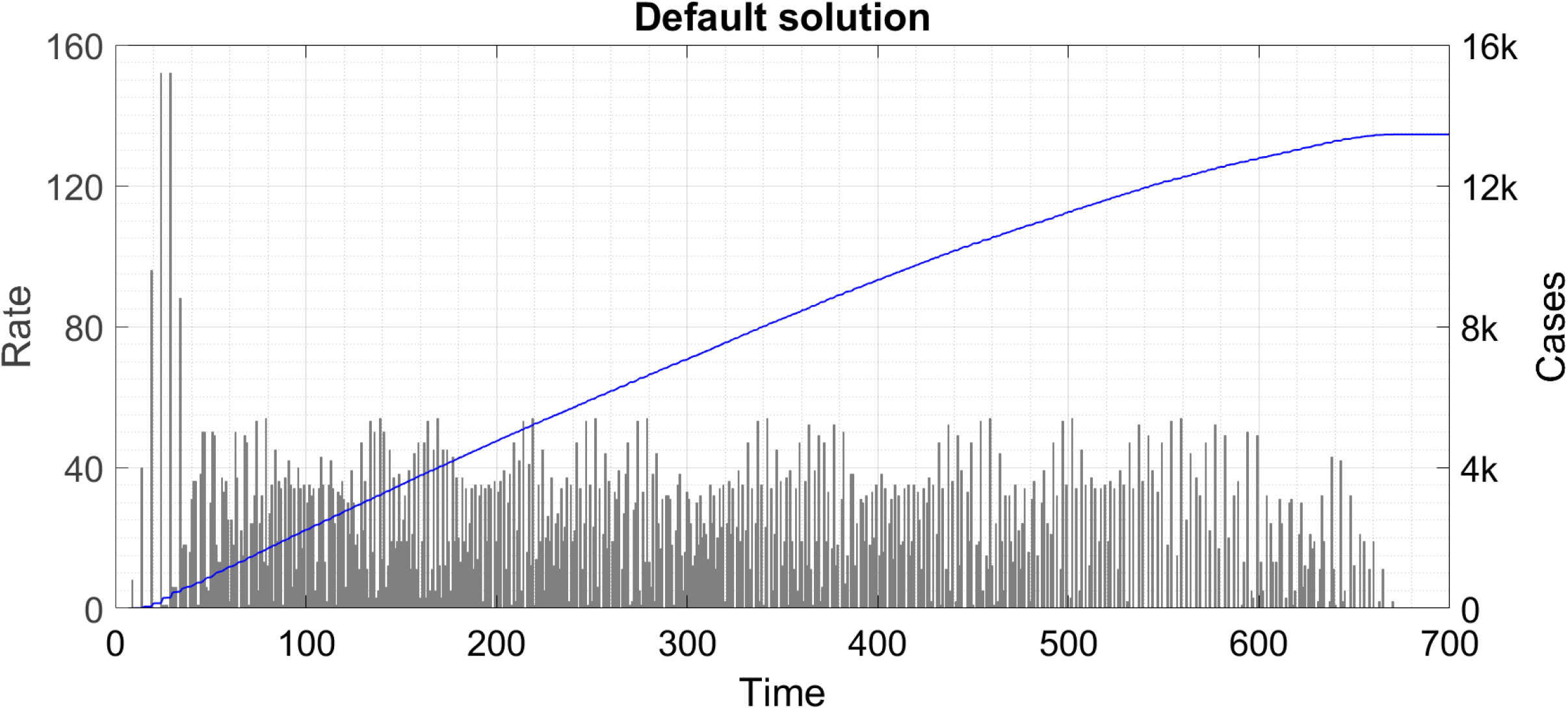
The default solution. The symbol ‘k’ denotes thousand.

**Figure 2:**
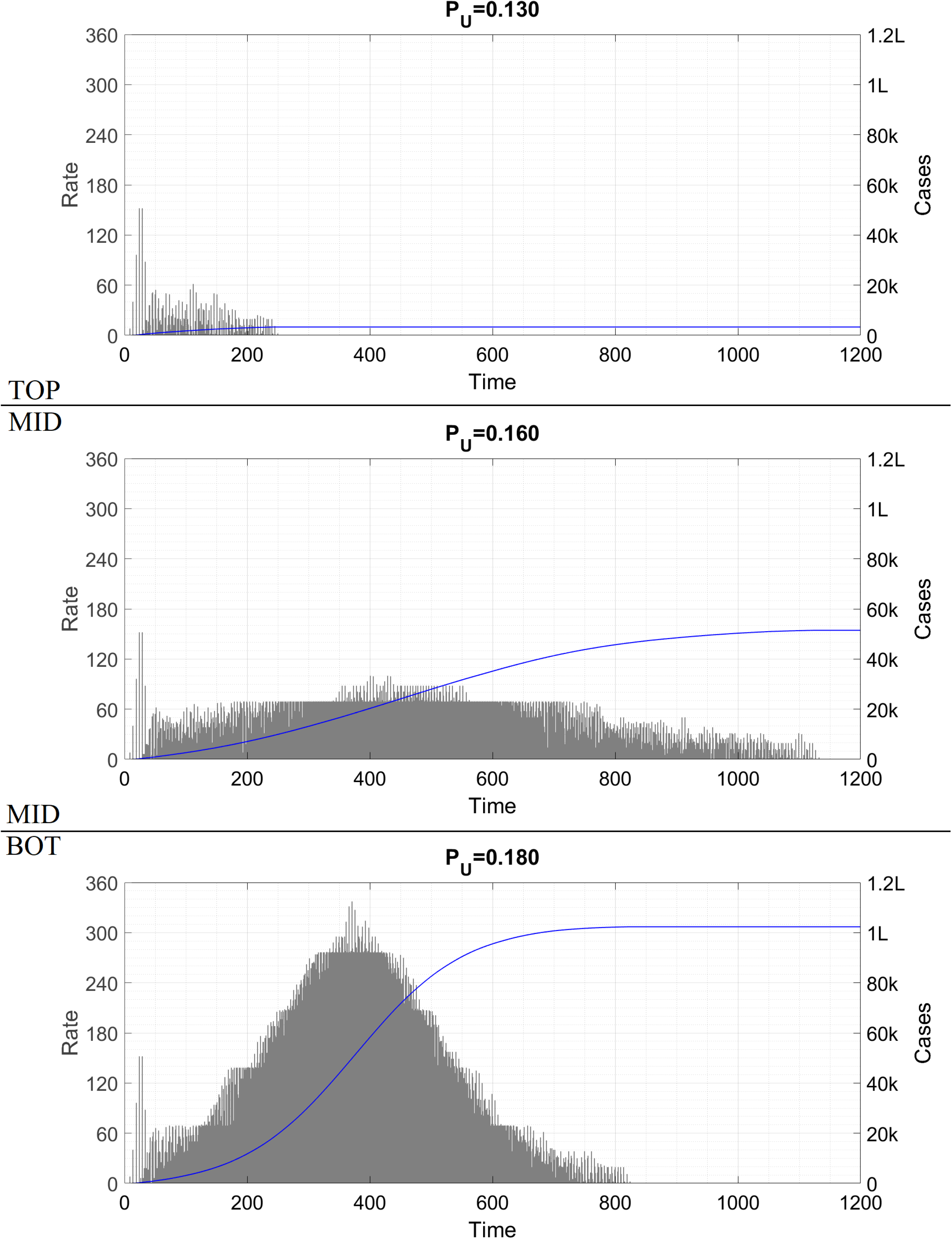
Time traces of the epidemic as P_U_ is varied. The symbol ‘k’ denotes thousand and ‘L’ hundred thousand.

**Figure 3:**
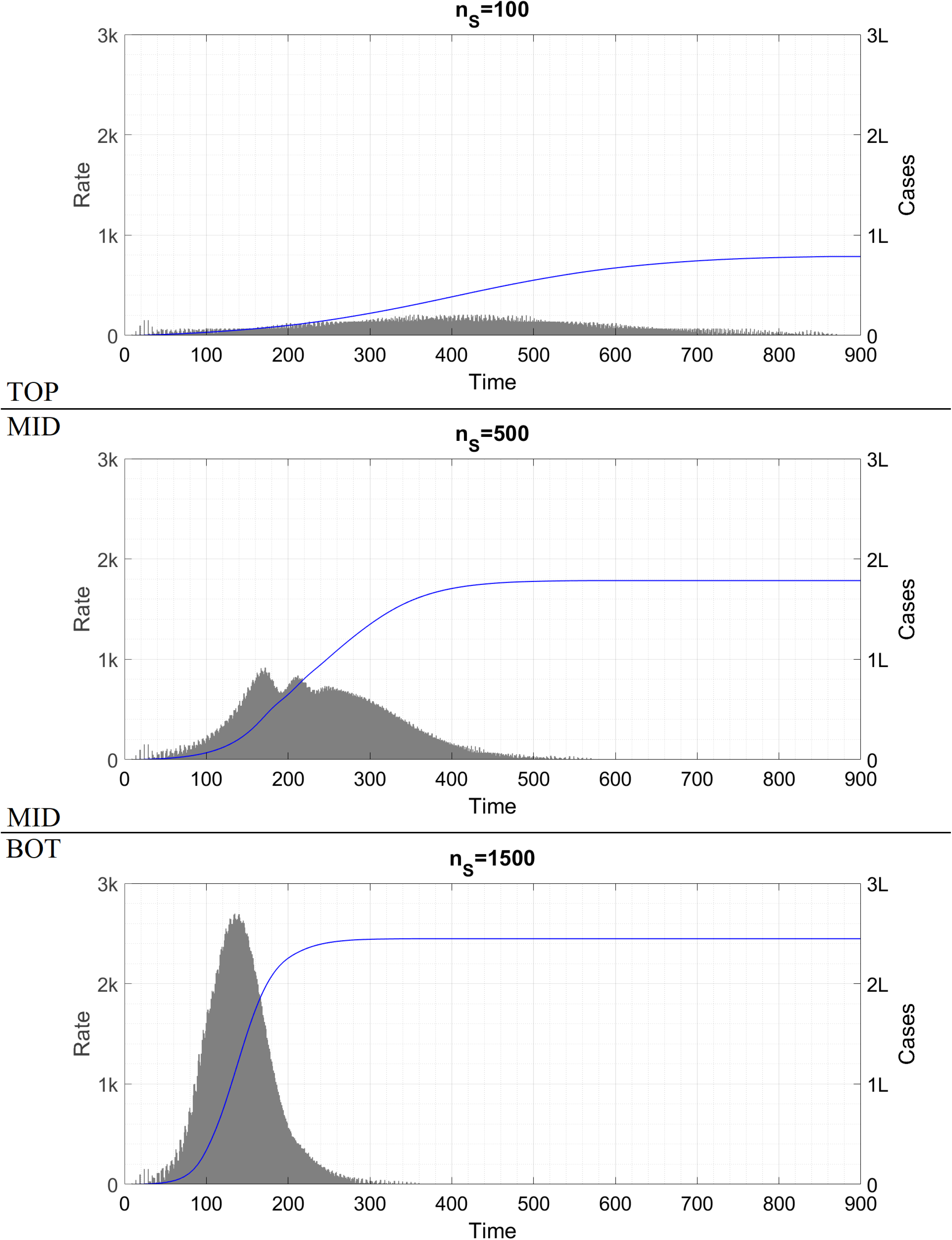
Time traces of the epidemic as n_S_ is varied. The symbol ‘k’ denotes thousand and ‘L’ hundred thousand.

We can see that for the smallest IC, not even one cluster is seeded and the epidemic stops at the imported cases. For the intermediate IC, six local clusters are seeded over 50 days before the epidemic runs out of steam. For the largest IC, the epidemic proceeds as we would expect it to. At all seeding vectors totalling 20 cases or more, we found the wave solution; below this threshold, it appeared that the boundaries were blurry. For example, the IC [4; 4; 4] actually led to a wave while [6; 6; 6] infected six clusters only. Similarly, nine initial cases could seed either zero local cluster or a few, depending on how they were distributed over the three days. To some extent, this variation might be the effect of the roundoff routine we are using – we shall clarify this in § 12.

As examples of the converse situation where a stable region is seeded with a very high caseload, we consider two scenarios in the Figure below. In the top panel, we take the default parameter values and seed it with a vector of external cases as above, but we choose this vector to be [120; 120; 120]. In the bottom panel, we start from the parameter values of Fig. 4 (default plus *n*_*S*_ = 1000) and seed it with the 8 initial clusters of Figs. 1-3. One hundred days into the epidemic however, we slash *n*_*S*_ to zero to bring all parameters to their default values.

**Figure 4:**
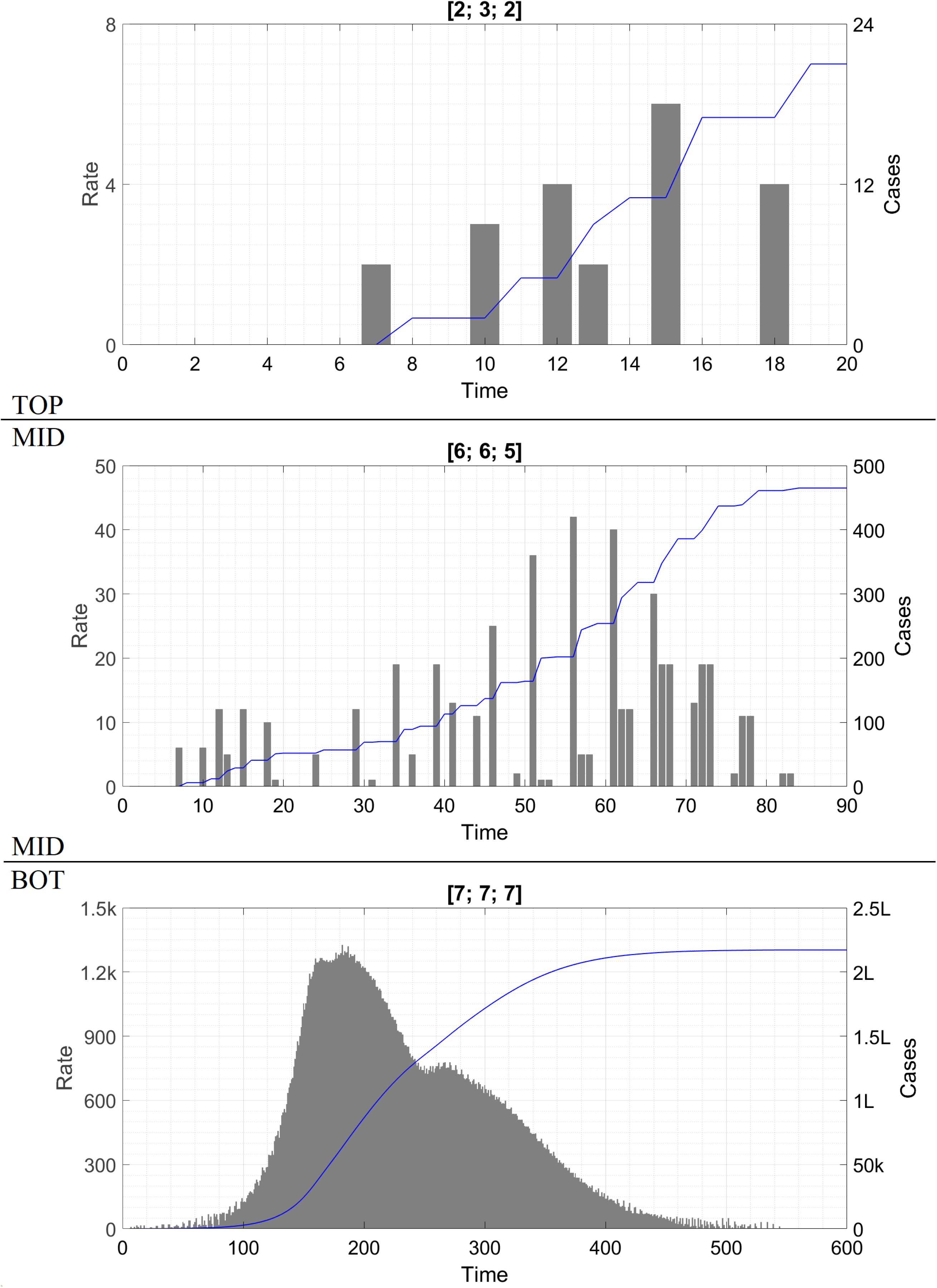
Time traces of the epidemic as the initial conditions are varied. The symbol ‘k’ denotes thousand and ‘L’ hundred thousand.

In the top panel, the cumulative caseload is higher than in Fig. 1 but the nature of the solution remains qualitatively unchanged. In the bottom panel, the cases decrease rapidly after the brakes are hit and the epidemic plateaus before petering out.

## DISCUSSION

### §10. Classical instability

The default plateau, Fig. 1, has first been reported by Thurner et. al. [2]. Figure 2 shows the effects of varying *P*_*U*_, the probability that an UCT event actually leads to a transmission (or equivalently, the average number of targets to whom one case transmits an infective dose of virus at an UCT event). *P*_*U*_ is governed by the degree of mask wearing, horizontal separation, hand-washing etc. It is a parameter which is closely related to classical epidemiological models – in the S-I-R model, it is accommodated into the force of infection while in the DDE model [1] it enters as a multiplicative factor in the per-case spreading rate (this is defined as *m*_0_ = *q*_0_*P*_0_ where *q*_0_ is the interaction rate and *P*_0_ the transmission probability). It is expected that a lower *P*_*U*_ will lead to a better outcome and vice versa, and Fig. 2 shows that this is indeed the case. Upto this point, our model agrees fully with classical models.

The first point of difference also comes up in Fig. 2. For, while at *P*_*U*_ = 0·130 the epidemic dies down in time after an initial phase of constant rate (probably caused by the strong seeding), at *P*_*U*_ = 0·160 the epidemic plateaus, just as it did with *P*_*U*_ = 0·150. Thus, the constant solution is actually valid for a range of *P*_*U*_ instead of just one value of *P*_*U*_ as happens in a classical model (the special value corresponding to *R*_0_ = 1). At sufficiently high *P*_*U*_ however, the constant solution is no longer seen, and is replaced by a wave solution. As we have already mentioned, the constant remains valid in the approximate range *P*_*U*_ belongs to [0·135, 0·165]. We shall say that *P*_*U*_ < 0·135 corresponds to a **super-stable** region of parameter space, 0·135 < *P*_*U*_ < 0·165 to a **stable** region of parameter space, and *P*_*U*_ > 0·165 to an **unstable** region of parameter space. Here, the usage of the words super-stable, stable and unstable is somewhat different from conventional dynamical systems theory, but this should not cause confusion. Within the stable range, the value of the constant rate and the duration of the epidemic both increase with increasing *P*_*U*_. In the unstable region, an increase in *P*_*U*_ causes the wave height to increase and the duration to decrease, in such a manner as to increase the cumulative caseload. This conclusion again agrees with the DDE model [1] and other classical models.

Note also that super-stable, stable and unstable regions of parameter space are all defined with respect to the prior infection level; for example, a parameter set which is unstable for fully susceptible population might be stable for 25 percent initial infection level and super-stable for 50 percent infection level. Thus, in the middle panel of Fig. 2, the mode of operation changes from stable to super-stable at approximately 800 days while in the bottom panel, the operation changes from unstable to stable/ super-stable at approximately 400 days (in the classical model, we say that *R* decreases across unity at this point). Practically, the existence of a constant solution over the entire stable range of parameter values complicates the epidemic management process as it implies that, seeing a linear solution in reality, we are not aware of how close we are to an instability. However, even after passing the instability, the system behaviour with increasing *P*_*U*_ remains tractable – a small increase in *P*_*U*_ causes only a gradual increase in case rate, which gives the authorities enough time to recognize the instability and reintroduce a higher level of NPI. This feature is again shared with the DDE model, where a small increase in *R* across the critical value of 1 causes a small increase in the case rate. Thus, Figs. 1-2 add no substantively new information beyond what can already be obtained from Refs. [1] and [2].

### §11. Cryptogenic instability

The instability in Fig. 3 however has no classical counterpart. We can see that even a small *n*_*S*_ of 100 can destabilize the plateau into a very broad and shallow wave – the cumulative caseload in Fig. 3-top is about six times higher than in Fig. 1. As *n*_*S*_ is further increased, the height and speed of the wave increase dramatically. What makes this instability even more surprising is that from the perspective of classical epidemiological models there is hardly any change at all in the interaction rate between the default solution and the three panels of Fig. 3. In these models, there is only a population-averaged spreading rate which is proportional to the average interaction or contact rate [14]. Let us calculate the average spreading rate for the situations here, when everyone is susceptible. In the default solution with no SEC events, each socially active case spreads the disease to 2 householders and approximately 2·5 cluster members (recall that the cluster sequence of [1; 3; 6; 7; 5; 1] is based on a very rough intra-cluster *R*_0_ of 2·5). In addition, this case participates in UCT on average once every 30 days (since *n*_*U*_/*N* is approximately 30) and spreads to an average of 0·15 person there. Thus, during three days (the transmissibility period as per our model), the case further spreads the disease to 0·15×(3/30) = 0·015 persons via UCT. Adding these two contributions, we can say that every socially active case spreads the disease to 4·515 persons or, more realistically, 4·5 persons. Since 1 in 3 cases are socially active and the others don’t spread at all, on average one case spreads the disease to 1·5 persons. (This is already a surprise since a classical *R*_0_ = 1·5 makes us expect a full-blown epidemic and not a constant crawl. But there is more to follow.) Dividing by the transmissibility duration gives us an average spreading rate of 0·5 person per day.

Now, consider the situation when *n*_*S*_ people participate in SEC events. By definition, each case present at these gatherings spreads the disease to 2 people, so the population-averaged spreading rate for SEC events is 2(*n*_*S*_/*N*) per day. For the three panels of Fig. 3, this works out to 0·00067, 0·0033 and 0·01 respectively. These increments are negligible relative to the 0·5 person per day contribution of household and intra-cluster (and UCT) transmission events. Classically, since the contact rate is proportional to the spreading rate, the SEC events add a negligible contribution to the contact rate. There is no way in which a 2 percent increase in contact rate can result in Fig. 1 being transformed into Fig. 3-bot. Hence, this instability does not exist in classical models and we call it **cryptogenic instability**.

Cryptogenic instability is dangerous from the viewpoint of epidemic management for two reasons. Firstly, unless we are aware of its existence, there is no reason to suspect that trips to restaurants and cinema halls constitute COVID-appropriate behaviour while attendance at wedding parties and birthday bashes does not. Secondly, unlike the classical instability where a small breach causes a small case growth, even a small breach here can cause a huge growth. By the time the authorities become aware of the danger and reimpose restrictions, the wave will already have overwhelmed healthcare systems.

The stark difference from classical formulations arises because in our model, the vast majority of a person’s interactions are confined within a small group of people (household and cluster) while the classical models assume that any person’s interactions are randomly and uniformly distributed among the entire population (this assumption is called homogeneous mixing). Thus, in our model, clusters start off explosively at *R*_0_ = 2·5, but they quickly turn ‘herd-immune’ and confine the bulk of the infection within themselves. In classical models, an outbreak with *R*_0_ = 2·5 can subside only after the entire population is herd-immune i.e. when nearly the entire region has been infected. The heterogeneity in interaction structure existing in the real world has been recognized by prior authors as well [15].

### §12. Critical mass effect

Figure 4 shows yet another phenomenon which is absent in classical epidemiological models. In these models, when the parameters are chosen to generate an instability (for example, in the DDE model [1] if the spreading rate *m*_0_ is taken above the critical value), even the smallest non-zero IC is sufficient to set off a wave of disease. Since the early growth is exponential, it hardly matters whether the IC features 100 cases or 1 case or 0·01 case (this latter being perfectly legitimate in a continuous model) – a small seeding can at most account for a delay in the peak by a couple of doubling times. Here however we see a huge difference. The parameters in Fig. 4 are chosen to lie in the highly unstable region, as the bottom panel demonstrates. Even so, it is possible for the outbreak to stop at just the seeding cases (top panel) or at the seeding cases plus a handful of clusters (middle panel).

We have already mentioned an anomaly regarding the ICs [4; 4; 4] vis-a-vis [6; 6; 6]. We believe that this is due to errors accumulated in the roundoff process – a lucky combination of values might infect a whole cluster and terminate the run while an unlucky combination might keep alive a fractional cluster which eventually adds up and perpetuates the epidemic. The presence of a discrepancy warrants a detailed analysis of the IC to verify that the effects shown in Fig. 4 are not spurious. There are two steps where rounding off takes place – once when calculating *γ* as rounded off *kP*_*U*_ and again when calculating the expectation value *E*(Δ*z*_*i*_). For really small numbers of cases, the concepts of roundoff or expectation value do not have too much meaning. A more relevant question is : given that there are *α* cases at large today, what is the probability that the virus is introduced into exactly 0, 1, 2 etc new clusters ? Since we are dealing with the start of the outbreak, we assume that all clusters are susceptible.

The bulk of the calculational framework we have already developed in § 4; a few extras needed to be taken care of. First is the probability that there are zero infected clusters – this eventuality was not relevant for calculating the expectation value as it would have had a null contribution. At SEC events, each case transmits to *m*_*S*_ = 2 people by definition, so zero transmission can occur if and only if zero cases are present at the events. The probability of this happening is

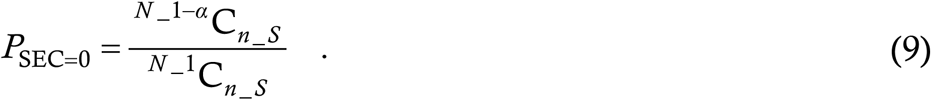

The calculation for *P*_UCT=0_ has one difference; while successful transmission to two people at SEC events is a certainty, transmission at UCT events is probabilistic with a chance of *P*_*U*_. From now onwards, we treat *P*_*U*_ as a genuine probability and not as an averaged quantity equivalent to *m*_*S*_, i.e. we assume that at each UCT event, a case transmits the disease to exactly one person with probability *P*_*U*_ and to zero person otherwise. Consequently, for *P*_UCT=0_, we must take into account not only the situation where there are no at large cases attending UCT events but also where there are *k* such cases and just none of them happen to transmit. The probability of there being *k* cases participating in UCT events has already been calculated in § 4; the probability that none transmit is (1 − *P*_*U*_)^*k*^. The probability of *k* = 0 is the exact equivalent of (9); taking this term and adding all the terms for *k* going from 1 to *α* yields

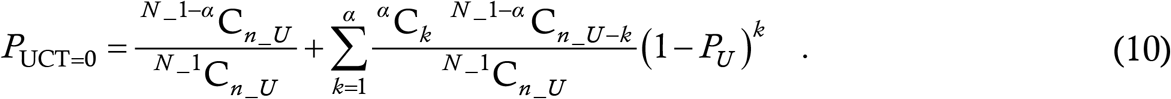

We now focus on infection of non-zero numbers of clusters.

The probability that SEC events infect exactly *b* clusters has already been calculated as part of (6); the expression is

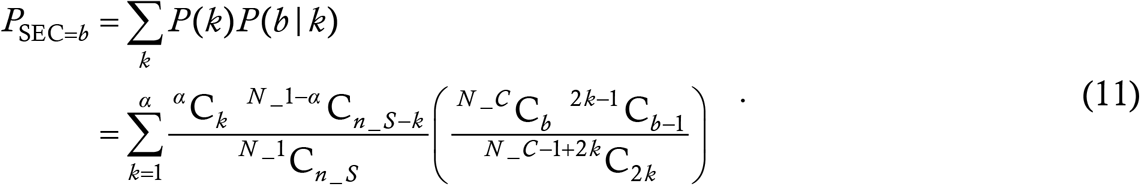

The probability that UCT events infect *b* clusters consists of three sub-probabilities, • that there are *k* cases at large, • that these *k* cases infect *j* new people with *j* going from 1 to *k*, and • that these *j* infectees belong to *b* clusters. The first and third of these probabilities have already been calculated in § 4; the second is identical to the probability that *k* tosses of a biased coin with probability *P*_*U*_ of heads result in *j* heads. This is ^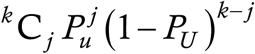^ to obtain *P*_UCT=_ *b* we must (as usual) multiply the sub-probabilities and sum over both *k* and *j* getting

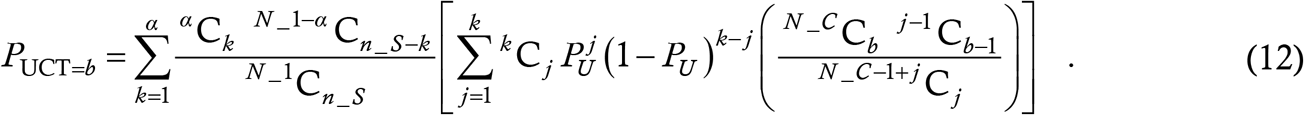

It is now a simple matter to compute the probability that *α* at large cases infect a total of *b* clusters. For *b* taking the values 0, 1 and 2 we have

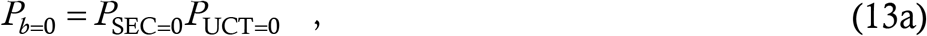

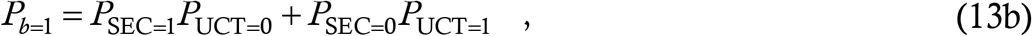

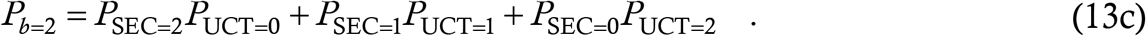

In the below Figure, we plot these probabilities as a function of *α* for the latter taking the values from 1 to 100. We use the parameter values of Fig. 4. We also plot 1 − *P*_*b*=0_ − *P*_*b*=1_ − *P*_*b*=2_, which is the probability that three or more clusters are infected.

This Figure shows that for low *α*, the likelihood of zero cluster’s being infected is very close to unity. As expected, this likelihood decreases as *α* increases but even at *α* = 28 the probability of no new infection is 1/2. Only at *α* = 51 does zero infection cease to be the most probable outcome. Thus, at low *α*, corresponding to a small external case influx or 1-2 infected clusters, it is indeed quite possible that the infection will not advance further in the population. This provides justification for the findings of Fig. 4.

Thus, even when a city is operating in the unstable region of parameter space, it needs a small but finite minimum number of initial cases to set the wave off. By analogy with nuclear reaction theory where a minimum quantity of fissile material is required to initiate the chain reaction, we call the seeding threshold the **critical mass**. From the viewpoint of epidemic management, the existence of critical mass is extremely dangerous. Seeded below this minimum, a city which is actually unstable will falsely behave like a super-stable or stable region, conveying a deceptive impression that the disease is under control when it is actually a disaster waiting to happen.

Figure 5 indicates that the converse of the critical mass phenomenon is not true – when the parameters are in the stable region, even a huge seeding cannot turn it unstable. We have checked this result for other parameter and seeding combinations as well and found it to be general. This is in agreement with the predictions of classical models where stability is independent of initial condition. It is a positive outcome from the viewpoint of epidemic management, since it implies that once tight controls are re-established following a surge, the presence of an existing huge number of cases will not cause the epidemic to propagate by itself like a perpetual motion machine.

**Figure 5:**
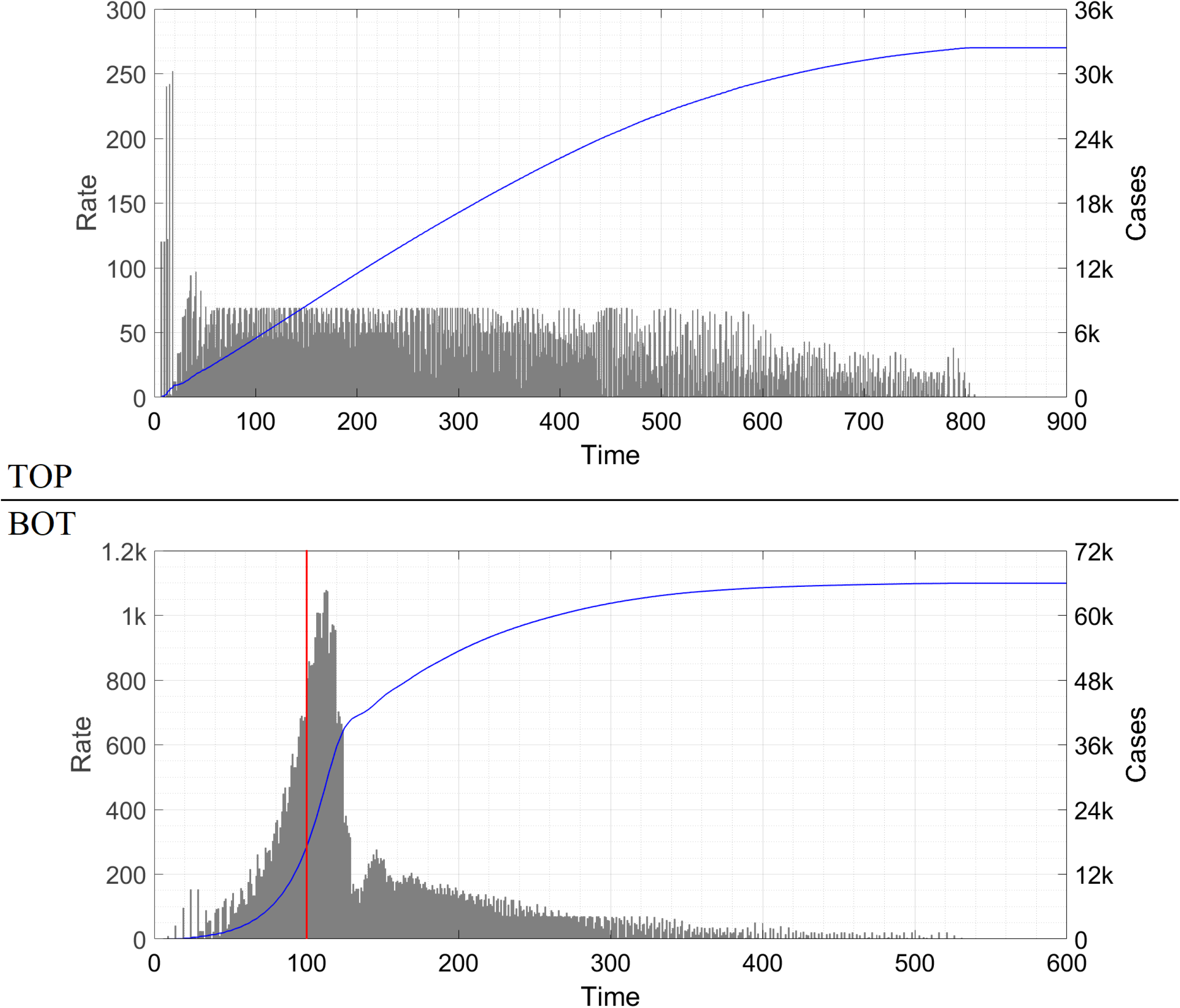
Time traces of the epidemic to demonstrate additional seeding-related effects. In the bottom panel the red line at day #100 indicates the abrupt reduction of n_S_ from 1000 to 0. The symbol ‘k’ denotes thousand.

**Figure 6:**
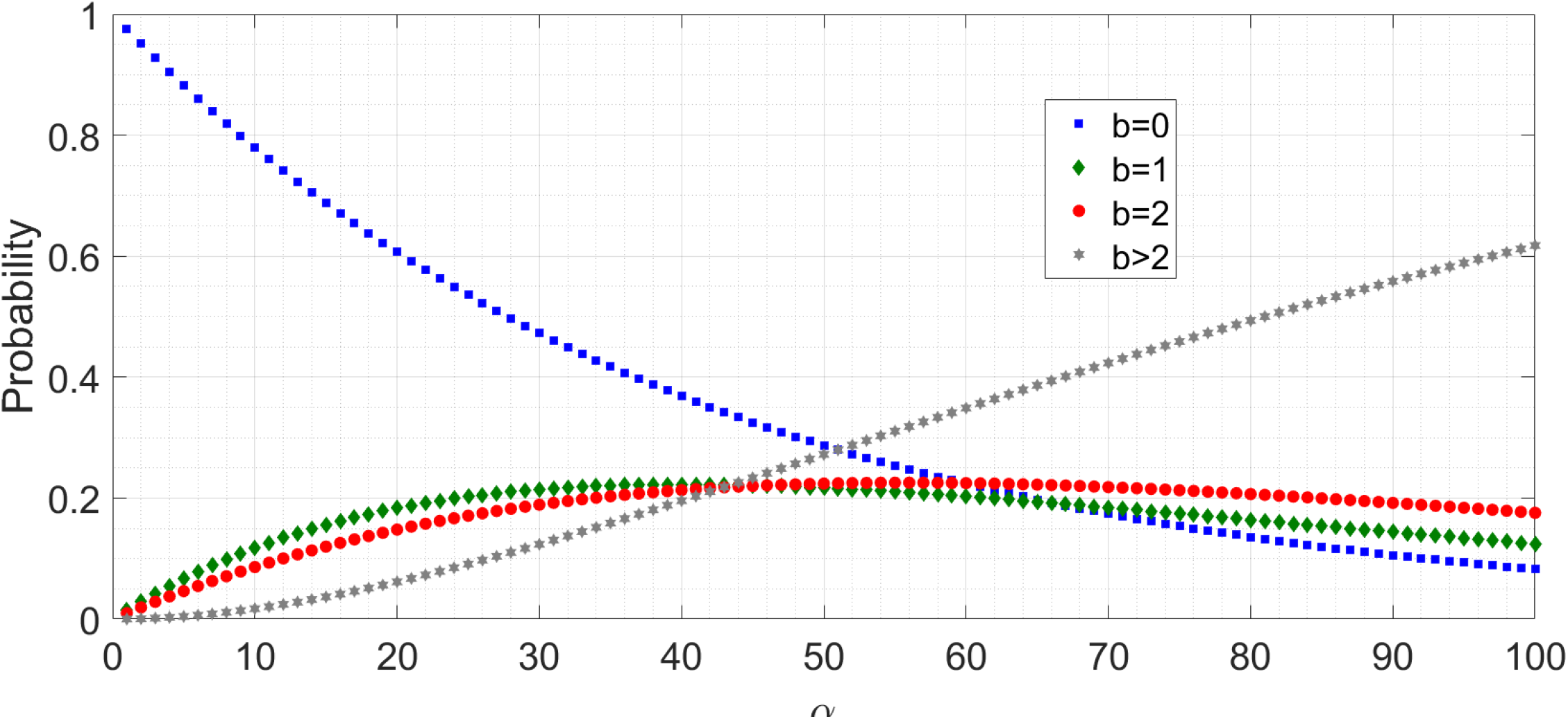
Probability of different number of clusters’ getting infected on any particular day, given the number of at large cases on that day.

While the general conclusions of Fig. 4 are robust, we do not set too great a store by Fig. 4-bot as an indicator of lockdown dynamics, especially if the lockdown is hard. This is because in a lockdown the clusters are forcibly broken up – groups of friends do not have the option to meet and socialize at entertainment venues. While case counts are expected to remain flat or even increase for a few days following lockdown on account of the serial interval and the household transmissions, the sharp increase seen in Fig. 4-bot between days #100 and #110 is less realistic as it arises from mechanically implementing the cluster sequence on all clusters seeded upto day #99. Similarly, the precipitous decline after day #110 and the dip near day #130 are also likely to be numerical artefacts. The rather slow decrease in case rate from day #150 onwards is however a most robust prediction – when case levels have fallen after a surge, we expect entertainment venues to be reopened which will resume intra-cluster transmission. In this regime, a persistent case level might be prevalent for a long time, and vaccination will be the best technique to drive it down.

### §13. Limitations and classical limit

Here we discuss the assumptions used in the model. Some obvious assumptions are those of constant household and cluster size. A more advanced formulation of the model can incorporate a distribution of household and cluster sizes. It can also account for the fact that more than one household member is socially active. Another assumption here is the constant cluster sequence of [1; 3; 6; 7; 5; 1], which remains valid even if multiple members of the same cluster are infected simultaneously. This sequence is a representation of an *R*_0_ = 2·5 dynamics in a large population, transferred to a small group. Accounting for multiple seeding will make the cluster get infected faster and thus bring forward a few cases by a few days; it will not affect the total count however. The basic phenomena we see here are independent of the specific choice of cluster sequence, but for a more accurate modeling we must use actual data collected from contact tracing activities (together with the necessary permissions). A caveat in the expressions (7) and (8) has already been mentioned in § 4 while the introduction of the parameter *k*_max_ has been discussed in § 5; these do not generate significant error.

Although the model incorporates a discrete population with heterogeneous interaction rates, it is eventually deterministic. This is necessary for computational tractability – a typical run with high caseload like one in Fig. 3 takes about five minutes on a laptop computer. For this purpose, we had no choice but to calculate an expectation value, with its rounding error as discussed in § 12. A more computationally advanced form of the model can be fully agent-based with every event assigned a certain probability. § 12 shows the beginnings of such an approach, where we calculated the probability of *α* cases infecting *b* clusters. A sophisticated computer might enable us to answer questions like “Given there are 100 cases today, what is the probability that there will be 1000 cases a month later ?” by calculating the probability of occurrence of every possible path from 100 to 1000 over 30 days (a daunting task by any standards). Such calculations can enable us to estimate best and worst case scenarios of the disease evolution corresponding to any given level of NPI.

Despite the approximations, our model is in excellent agreement with reality. Just as a physical theory is eventually vetted by agreement with experiment, so too the ultimate validation of our model is its explanation of the widely seen plateau states and their stability transitions to epidemic waves. In addition, our model relies on hypotheses – primarily that of cluster-based interaction – which are plausible on an absolute scale.

In addition, we now demonstrate that our model reduces to the classical model in the appropriate limit. To incorporate homogeneous mixing (the unstated or at best understated pillar of classical models), we consider the situation where households are scrapped (so that *N* = *N*_1_) and clusters are reduced to size unity (*N*_*C*_ = *N, y*_*i*_ = *z*_*i*_ and Δ*y*_*i*_ = Δ*z*_*i*_ for all *i*). In this limit, UCT and SEC modes of transmission are equivalent; let us merge them into the SEC category. *m*_*S*_ remains the number of people to whom one case spreads the disease in a day if they are all susceptible. To incorporate continuous mixing (another classical assumption) we let *n*_*S*_ = *N* i.e. we allow the entire population to mix every day. Under these assumptions let us run through Algorithm 1.

In a classical model, we do not need rounding off so we discard the subroutine roundoff. In the main routine, everything upto the primary loop over *i* remains unchanged. In Step 1 of the primary loop we let go of *k*_max_. In Step 2, (6) as it stands no longer has meaning. Since everyone participates in the ‘SEC’ events, *k* can take only the value *α* – the first summation and the expression for *P* (*k*) both vanish. The number of people who receive the infective dose of virus remains *β* = *m*_*s*_*α*. By definition these *β* people belong to exactly *β* ‘clusters’ so the second summation in (6) is redundant as well. What remains is the third summation i.e. the calculation of the expected number of susceptible clusters which are seeded (or equivalently the expected number of susceptible persons who are infected).

The problem we have on hand is, given that there are *N* − *y*_*i*_ people who are susceptible and *y*_*i*_ people who are immune, what is the probability *P* (*j*) that among *β* randomly selected people who receive an infective dose, exactly *j* are susceptible ? Then the expectation value can be calculated as *E* = Σ _*j*_ *jP* (*j*) as previously, with *j* running from 1 to *β*. The fully accurate expression for *P* (*j*) is

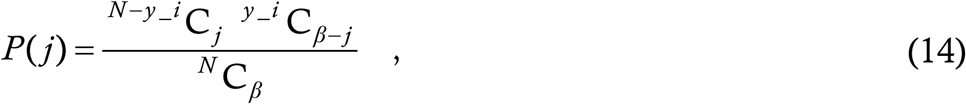

as we have already calculated several times in different contexts. However, if *β* << *y*_*i*_, *N*−*y*_*i*_, i.e. the daily new cases are very low compared to the susceptible and immune populations (a very reasonable assumption even in the worst of worst-hit areas unless we are extremely close to the beginning of the epidemic), then we can assume that each individual person is susceptible with probability *p* = 1 − *y*_*i*_/*N* and immune with probability 1− *p*. (An equivalent problem is that a box contains *y*_*i*_ blue balls and *N* − *y*_*i*_ green balls; if *β* balls are drawn at random without replacement, then what is the probability that *j* balls are green ? In the limit *y*_*i*_, *N*−*y*_*i*_ >> *β*, we can replace this by the situation where the balls are drawn *with* replacement.) With this assumption, *P* (*j*) becomes

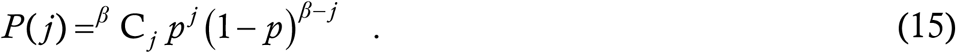

The expectation value of a binomial distribution is a standard formula in probability theory [16]; in our problem it evaluates to *βp* = *β* (1 − *y*_*i*_/*N*). Thus, in Step 2 of Algorithm 1, instead of the expression (6) we now use the value *E*_*S*_ = *β* (1 − *y*_*i*_/*N*).

Since UCT events have been merged with SEC events and since rounding off is no longer significant, we skip Steps 3 to 6. Step 7 remains as is with **v** being the singleton vector [1] (not Reference 1) and *h* taking the value 1. So, putting everything together and using the values of *α* and *β*, we have

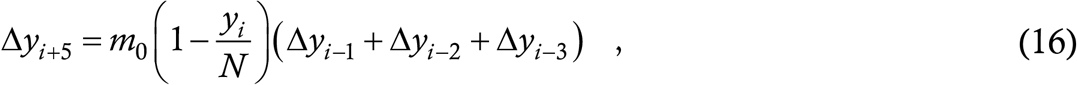

where, since SEC and UCT events have lost their separate identities, *m*_0_ is more appropriate than *m*_*S*_. We now want to express this as a continuous time system or flow rather than a map, so let the continuous function *y* (*t*) denote the cumulative count of corona cases as a function of time. Step 8 of Algorithm 1 tells us that Δ*y*_*i*_ is the equivalent of d*y*/d*t*; the big bracket in the right hand side of (16) above is the number of new cases which have cropped up over the last three days, which is *y* (*t*) − *y* (*t* −3). Using this we have

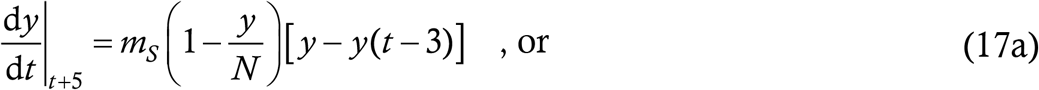

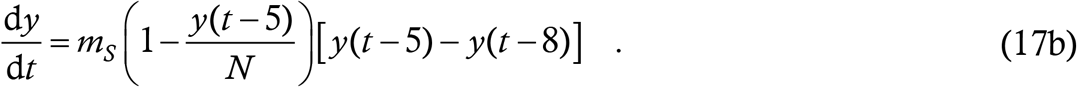

Equation (17b) agrees exactly with Equation (29) of Ref. [1]; if we exclude the 5-day delay which merely shifts the infection curve rightwards by 5 days, it becomes the standard retarded logistic equation. Thus, the classical model emerges as a limiting case of the present model.

Although the DDE (17b) is a special case of Algorithm 1, and although Algorithm 1 is capable of predictions which (17b) is not, there are nonetheless some occasions where (17b) is more useful than Algorithm 1. For example, the averaging assumptions allow us to smoothly accommodate asymptomatic and symptomatic cases who have different transmissibility durations. This is much harder to implement in this model. A detailed theory of contact tracing can be built into the DDE model [1] which is much more difficult here – a contact trace is expected to break transmission partway through a cluster, but what happens to the susceptible members after the traced cases recover ? Cluster fragmentation is currently not incorporated into this model. Age-structuring is another phenomenon which is easier to model using differential equations. Finally, as mentioned in § 12, rigorous lockdowns (full lock or at least closure of entertainment venues) are likely to drastically reduce the cluster size or make the cluster concept irrelevant altogether. In such a situation, the socially active people are expected to behave like millions of tiny clusters, which is better described by (17b) than by Algorithm 1.

### §14. Conclusion

In this Article we have proposed a mathematical model for the spread of COVID-19 which takes into account the heterogeneity in human interaction patterns. This model yields a plateau as a generic solution and also demonstrates the pathways by which it can be destabilized into a mountain. In future, we shall analyse how the phenomena outlined here lead to an understanding of the case trajectories in India as well as other countries. While COVID-19 remains our immediate focus, the concepts we have presented are valid for any infectious disease. Hence, our model and findings should have utility in the management of future epidemics as well.

## DECLARATIONS

We have NO conflict of interest to declare.

We have NOT been funded by any agency, public or private, for this study.

## Data Availability

All data used here are available on internet websites which we have cited

## APPENDIX

### §A1. Computer evaluation of (5), (6)

While running the simulations, we observed that evaluation of the expressions (5) and (6) was impossible for the computer unless they were inputted in a special form. This happened because each of the probabilities *P* (*k*), *P* (*b*| *k*) and *P* (*j*| *b*) features a ratio of two huge numbers, both of which are beyond the computer’s calculational capacity. However, each huge number here is a product of many smaller numbers, and the computer has no trouble handling these individually. Thus, in the expression

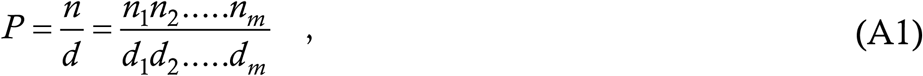

*n* and *d* are beyond the computer’s resources while *n*_1_, *n*_2_, *d*_1_, *d*_2_ etc are not. To evaluate *P*, we must enter it as (*n*_1_/*d*_1_) × (*n*_2_/*d*_2_) × ….. ×(*n*_*m*_/*d*_*m*_) with *n*_*i*_ and *d*_*i*_ preferably being of comparable size, and then the computer has no difficulty in the calculation. Analytically opening out the combinations, we have written the probabilities *P* (*k*), *P* (*b*| *k*) and *P* (*j*| *b*) for easy machine evaluation in the forms described below.

The analytical formula for *P* (*k*) is

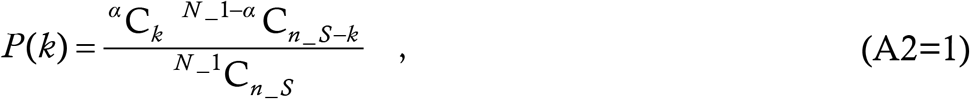

where we have repeated (1) of the main text as (A2) here just for display convenience. For the computer we define this as a function g = prob1(a,k,N,s) where N denotes *N*_1_ and s denotes *n*_*S*_. Opening out the combinations and rearranging items so that large numerators carry large denominators (note that N is the really large number here while the others are much smaller), we find

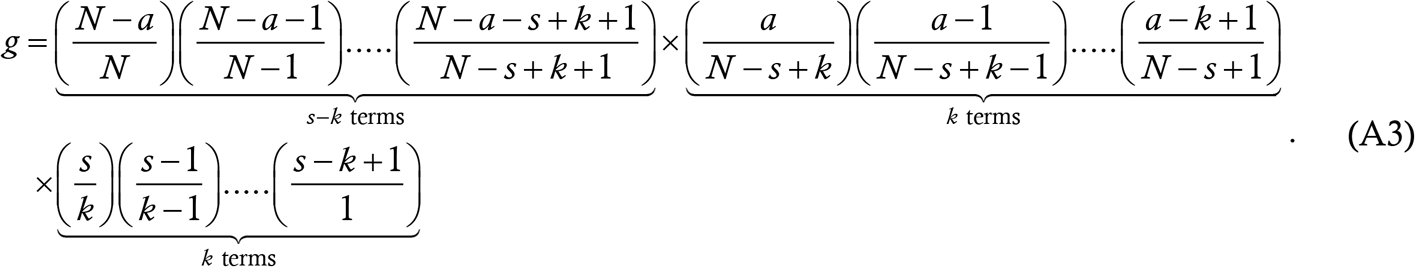

Since k cannot be zero and is likely to be less than s (it is extremely improbable that every single person attending SEC gets infected), there are no borderline cases to be taken care of manually.

The analytical formula for *P* (*b*| *β*) is

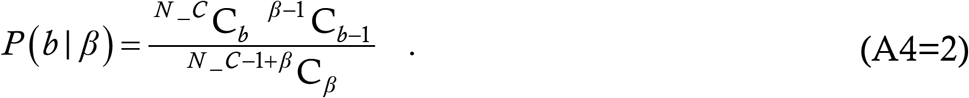

For the computer we define the function g = prob2(N,b,t) where N denotes *N*_*C*_ and t denotes *β*. This function is

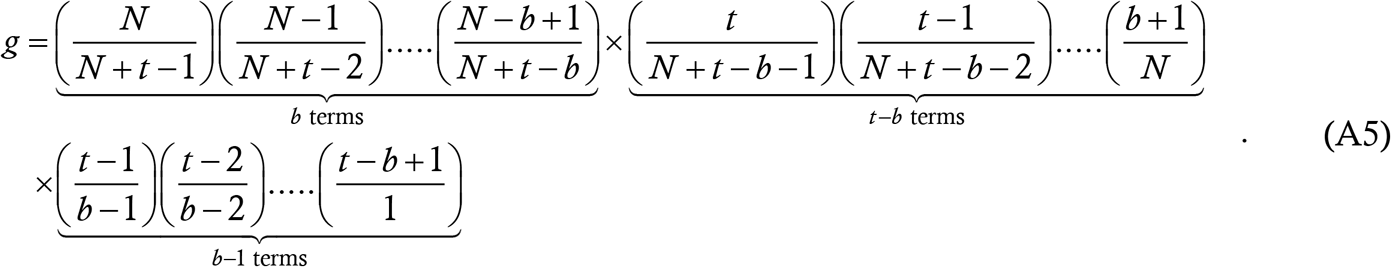

Unlike with (A3), this has marginal cases. If t = 1 (impossible for SEC where *β* is defined as 2*k* but quite possible for UCT) then b must equal 1 and g must be unity. If b = t (a physically meaningful case) then the second product in (A5) is non-existent i.e. it must equal unity. If b = 1 (for whatever t) then the third product must be bypassed and set to unity.

The analytical formula for *P* (*j*| *b*) is

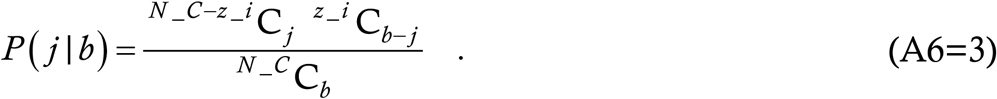

For the computer we define the function g = prob3(z,j,N,b) where z denotes *z*_*i*_ and N denotes *N*_*C*_. We recognize that this is the same as the function g = prob1(z,b-j,N,b) except for the boundary cases which we recognize by looking at (A3). If b = j then g can be evaluated manually as

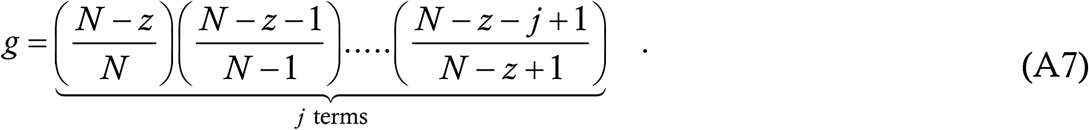

If z = N i.e. all clusters have been infected then g = 0. In all other circumstances, we define q = b-j and set g = prob1(z,q,N,b).

